# Analysis of adaptive immune cell populations and phenotypes in the patients infected by SARS-CoV-2

**DOI:** 10.1101/2020.03.23.20040675

**Authors:** Xiaofeng Yang, Tongxin Dai, Xiaobo Zhou, Hongbo Qian, Rui Guo, Lei Lei, Xingzhe Zhang, Dan Zhang, Lin Shi, Yanbin Cheng, Jinsong Hu, Yaling Guo, Baojun Zhang

## Abstract

Coronavirus disease-2019 (COVID-19), caused by SARS-CoV-2, has rapidly spread to most of countries in the world, threatening the health and lives of many people. Unfortunately, information regarding the immunological characteristics in COVID-19 patients remains limited. Here we collected the blood samples from 18 healthy donors (HD) and 38 COVID-19 patients to analyze changes in the adaptive immune cell populations and phenotypes. In comparison to HD, the lymphocyte percentage was slightly decreased, the percentages of CD4 and CD8 T cells in lymphocytes are similar, whereas B cell percentage increased in COVID-19 patients. T cells, especially CD8 T cells, showed an enhanced expression of late activation marker CD25 and exhaustion marker PD-1. Importantly, SARS-CoV-2 induced an increased percentage of T follicular helpher (Tfh)- and germinal center B-like (GCB-like) cells in the blood. However, the parameters in COVD-19 patients remained unchanged across various age groups. Therefore, we demonstrated that the T and B cells can be activated normally and exhibit functional features. These data provide a clue that the adaptive immunity in most people could be primed to induce a significant immune response against SARS-CoV-2 infection upon receiving standard medical care.

---

A severe pneumonia-associated respiratory syndrome began in Wuhan of China in December 2019, calling the attention of WHO, which subsequently declared the disease as a public health emergency of international concern. The novel coronavirus strain was officially named as severe acute respiratory syndrome coronavirus 2 (SARS-CoV-2) [1, 2]. Coronaviruses infections such as severe acute respiratory syndrome (SARS) and Middle East respiratory syndrome (MERS), can cause severe respiratory disease [3, 4]. SARS-CoV-2 is an enveloped positive-strand RNA virus, which belongs to the family of coronaviruses along with SARS-CoV and MERS-CoV according to the genome similarity [2, 5-7].

A number of studies demonstrated that the adaptive immunity responds to coronavirus and is required for efficient clearance of the virus. In patients infected with SARS-CoV, the acute phase of infection in humans was associated with a severe reduction of T cell numbers in the blood, involving a dramatic loss of CD4 and CD8 T cells in comparison to healthy control individuals [8, 9]. This suggests that SARS-CoV infection impaired cellular immunity in the early stages of the disease. With the prolonged recovery time of SARS-infected patients, activated T cell markers such as CD69 and CD25 expression decreased[10, 11], indicating that T cell activation in response to the virus is impaired [12]. With the improvement of the disease, the ratio of CD4 to CD8 T cells increased, indicating that CD4 T cells recovered faster than CD8 T cells [13]. In addition, within the 92 % of cured SARS patients whose B cells declined first and then increased or continued to increase during the course of the disease, only 8 % of them had a constant or decreasing cell count [14]. Similar to SARS infection in humans, leukopenia and lymphopenia are also observed in MERS patients, albeit to a lesser degree than that observed in SARS patients. A detailed clinical study showed that 14% of MERS patients were leukopenic while 34% of the patients had lymphopenia[15]. MERS-CoV-infected patients that exhibited distinctively high frequencies of MERS coronavirus–reactive CD8 T cells were associated with severe/moderate illness, whereas CD4 T cell response was minimally detected at this stage. In the convalescent phase, slightly more CD4 T cells were detected [15].

Currently, very few studies showed that COVID-19 patients underwent developing lymphopenia and rising pro-inflammatory cytokines in severe cases [16-18]. The information is still very limited about how immune cells change and function in response to SARS-COV-2 infection. Knowing that T and B cells respond to the infection and play critical roles in defending against virus infection, systematically studying the changes in T and B cells in COVID-19 patients will help uncover the immune response against SARS-COV-2 infection and provide insights for COVID-19 diagnosis and treatment.

In this study, we analyzed the blood samples from 18 HD and 38 patients and focused on the characterization of adaptive immune cell populations and phenotypes upon SARS-CoV-2 infection. We showed that upon infection, lymphocyte percentage declined, the percentages of CD4 and CD8 T cells within the lymphocyte population remained unchanged, and B cell percentage was relatively increased. CD4 and CD8 T cells exhibited a mild and strong activation phenotype, respectively. Notably, the percentages of Tfh- and GCB-like cells increased. Similar phenotypes among the patients in various age groups indicate that aged individuals are also capable to respond to SARS-CoV-2 infection. Our data support the notion that adaptive immunity could be normally activated and defend against SARS-CoV-2 infection.

## Results

### The percentage analysis of T and B cells in COVID-19 patients

To determine the change in the composition of adaptive immune cells, we analyzed the percentages of T and B cells in the blood from HD and patients using flow cytometry. In comparison to HD, lymphocyte percentage in the whole blood was not significantly changed, though exhibited a decreasing trend in patients (Fig.1A). Within the lymphocyte population, the percentages of CD4^+^ and CD8^+^ T cells were comparable (Fig.1B and C), whereas B cell percentage significantly increased (Fig.1D) in COVID-19 patients.

**Fig 1.**
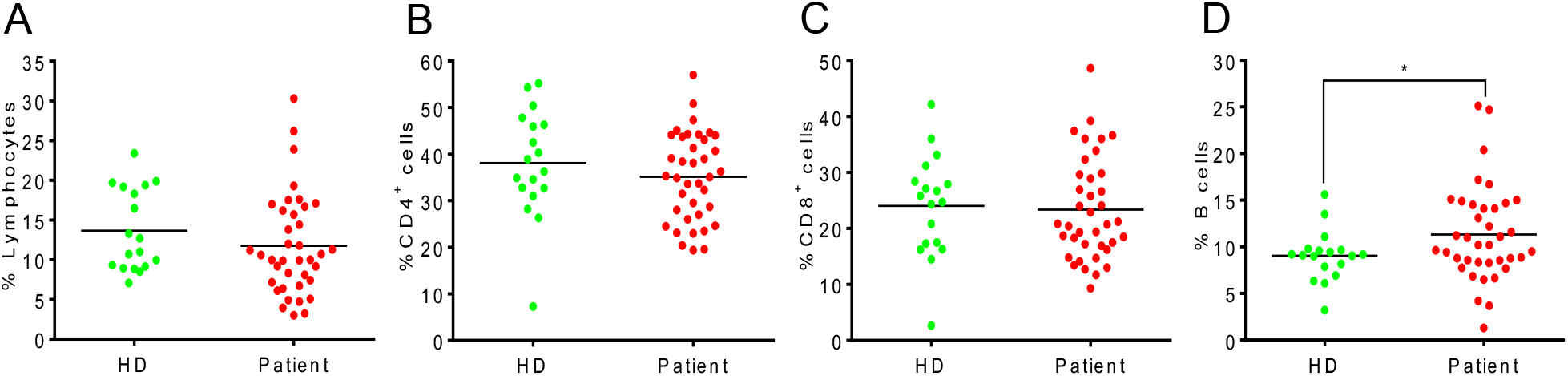
The percentage changes of T and B cells between HD and COVID-19 patients. (A)The percentage of lymphocytes in total blood cells. (B) The percentage of CD4^+^ T cells in lymphocyte population. (C) The percentage of CD8^+^ T cells in lymphocyte population. (D) The percentage of B cells in lymphocyte population. Each dot represents a single patient of COVID-19 or healthy donor. * P<0.05 was considered statistically significant.

### An activated phenotype of T cells in COVID-19 patients

To evaluate T cell status in response to SARS-CoV-2 infection, we analyzed the expression of CD69, CD25, PD-1, CD45RA, CD45RO and CXCR3 in both CD4^+^ and CD8^+^ T cells. In CD4^+^ T cells of COVID-19 patients, the expression of CD69 and CD25 (Fig. 2A and B) and the percentage of regulatory T cells (Tregs), marked by CD3^+^CD4^+^CD25^+^CD127^-^ (Fig. 2F) were similar to that of HD. CD25 expression upregulated significantly in CD8^+^ T cell population of the patients (Fig. 3B). The proportion of naïve and effector/memory cells in both CD4^+^ T cells (Fig. 2D and E) and CD8^+^ T cells (Fig. 2E and F) of the two groups were not significantly different. PD-1 expression upregulated dramatically in both CD4^+^ T cells (Fig. 2C) and CD8^+^ T cells (Fig. 3C) of the patients. The data demonstrated a weak activation in CD4^+^ T cells, but a strong activation in CD8^+^ T cells during SARS-CoV-2 infection.

**Fig 2.**
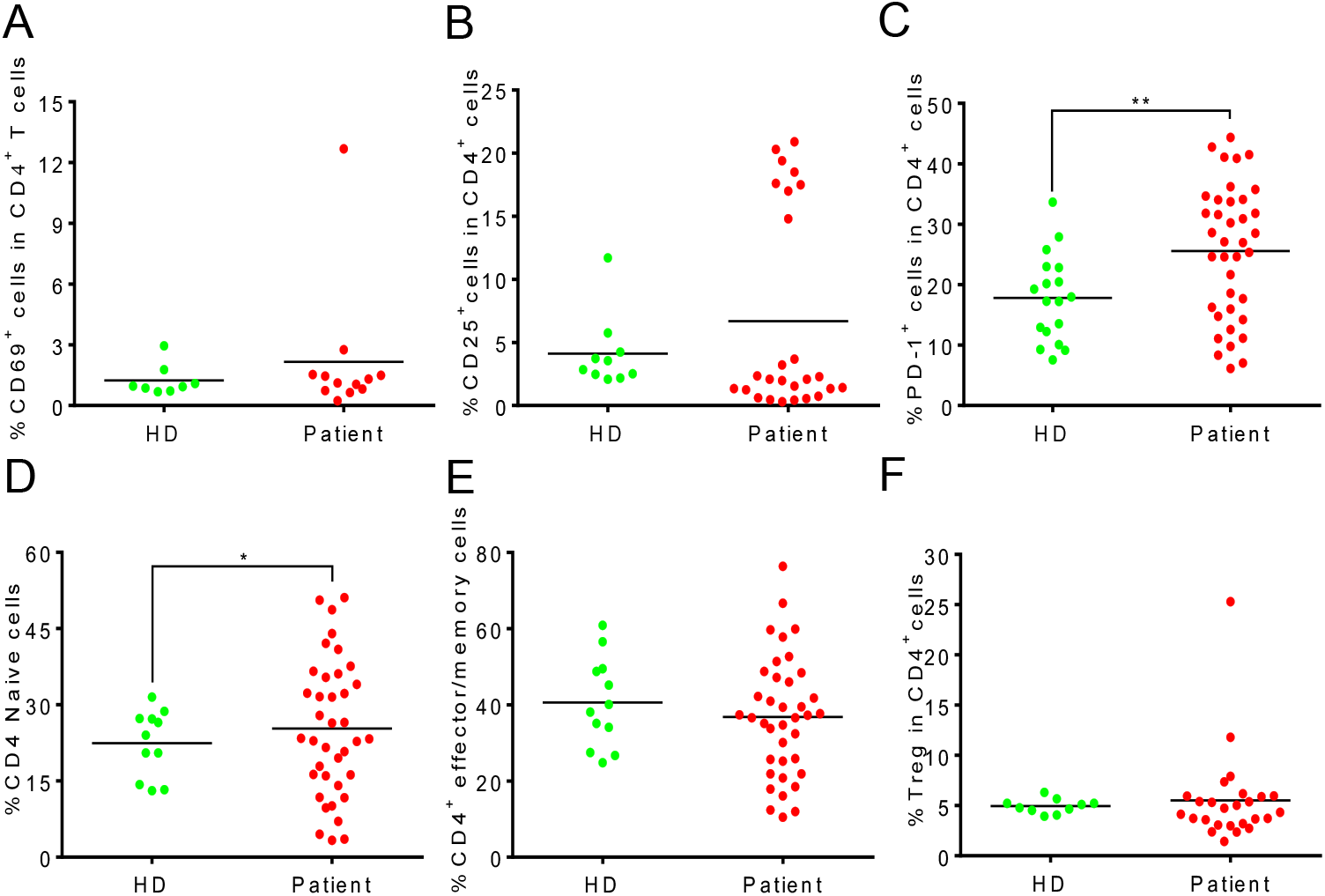
A slight increase of activated CD4+ T cells in COVID-19 patients. (A)The percentage of CD69^+^ cells in CD4^+^ T cells. (B) The percentage of CD25^+^ cells in CD4^+^ T cells. (C) The percentage of PD-1^+^ cells in CD4^+^ T cells. (D) The percentage of CD45RA^+^CD45RO^-^ cells in CD4^+^ T cells. (E) The percentage of CD45RA^-^CD45RO^+^ cells in CD4^+^ T cells. (F) The percentage of Treg cells in CD4^+^ T cells. Each dot represents a single patient of COVID-19 or healthy donor. * P<0.05 and P<0.01 was considered statistically significant and extremely significant, respectively.

**Fig 3.**
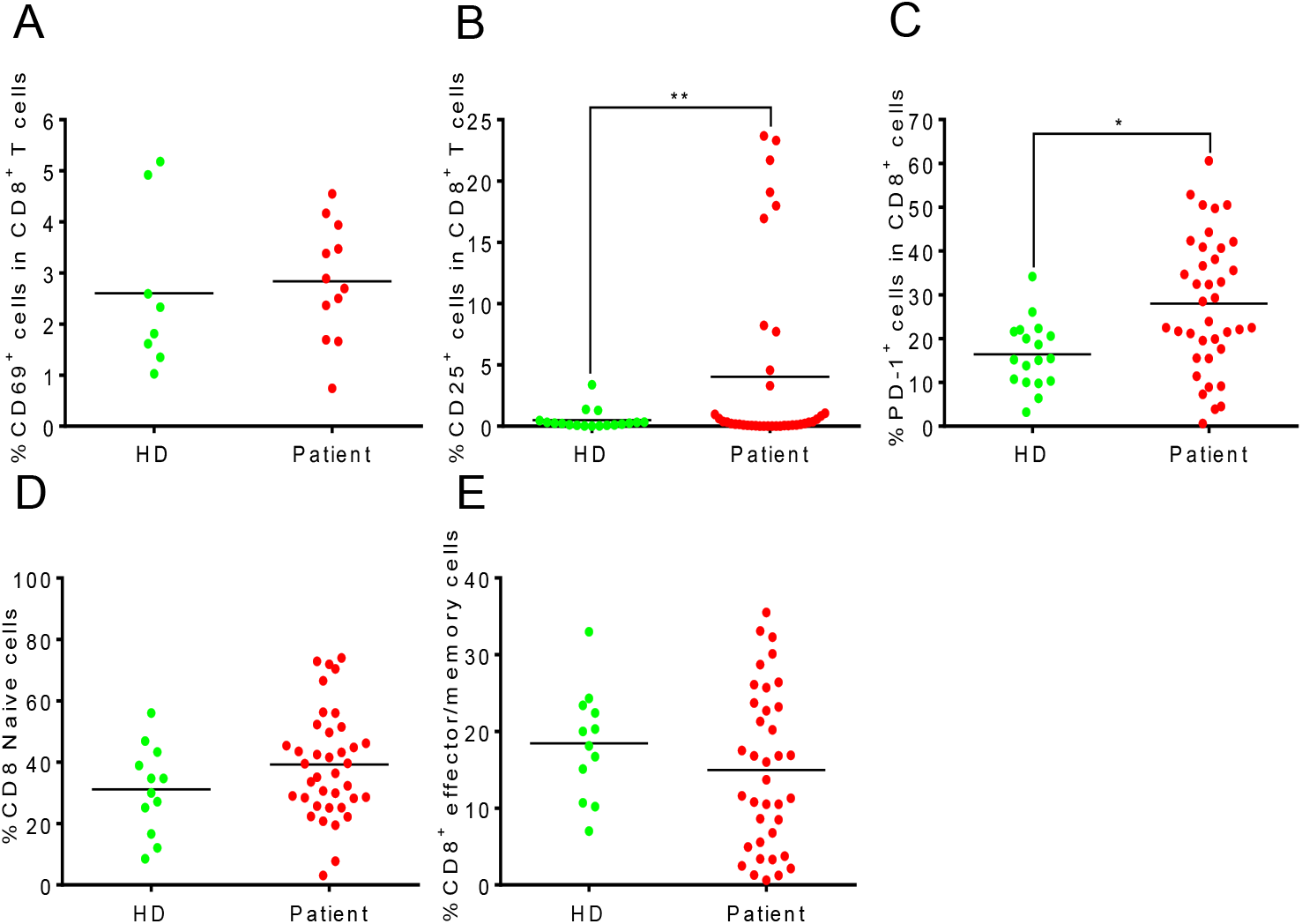
A strong increase of activated CD8+ T cells in COVID-19 patients. (A)The percentage of CD69^+^ cells in CD8^+^ T cells. (B) The percentage of CD25^+^ cells in CD8^+^ T cells. (C) The percentage of PD-1^+^ cells in CD8^+^ T cells. (D) The percentage of CD45RA^+^CD45RO^-^ cells in CD8^+^ T cells. (E) The percentage of CD45RA^-^CD45RO^+^ cells in CD8^+^ T cells. Each dot represents a single patient of COVID-19 or healthy donor. * P<0.05 and P<0.01 was considered statistically significant and extremely significant, respectively.

### An increase in germinal center-like cells in COVID-19 patients

T follicular helper (Tfh) cells can help B cell activate and differentiate into effector cells, produce high-affinity antibody and form germinal centers (GC)[19]. To study whether COVID-19 patients produce efficient adaptive immune response, we analyzed the expression of PD-1 and CXCR5 in CD4^+^ T cells, and the expression of Fas and GL7 in B cells. As shown in Figure 4A and B, there was a significant increase of both Tfh-and GCB-like cells in the blood of the patients compared to HD group.

**Fig 4.**
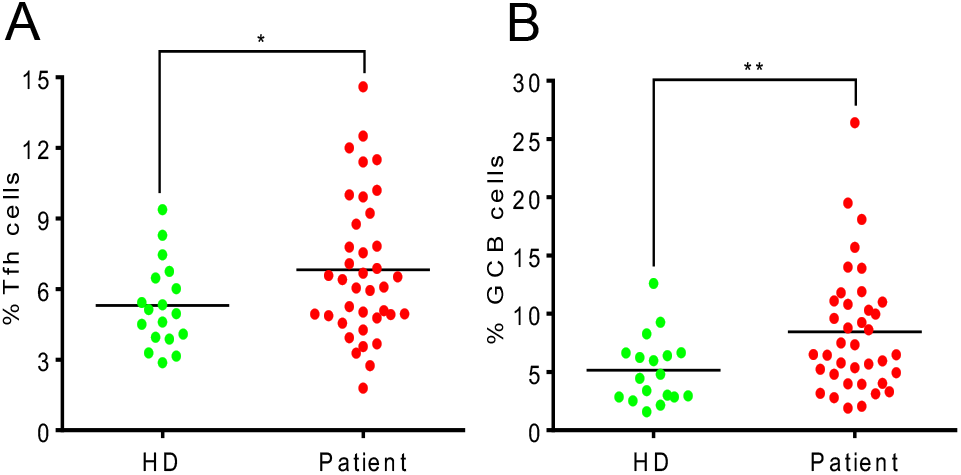
An increase of germinal center-like cells in COVID-19 patients. (A)The percentage of PD-1^+^CXCR5^+^ cells in CD4^+^ T cells. (B) The percentage of Fas^+^GL7^+^ cells in B cells. Each dot represents a single patient of COVID-19 or healthy donor. * P<0.05 and P<0.01 was considered statistically significant and extremely significant, respectively.

### Correlation analysis between activation signature and patient age

To study whether age affects adaptive immune cell population and effector features, we performed correlation analysis between T cell activation markers and age. In Figure 5, no dramatic change occurred with increasing age. The result indicates that in the aged individuals infected by SARS-CoV-2, there is no defect in CD8^+^ T cell activation, as well as Tfh- and GCB-like cell differentiation.

**Fig 5.**
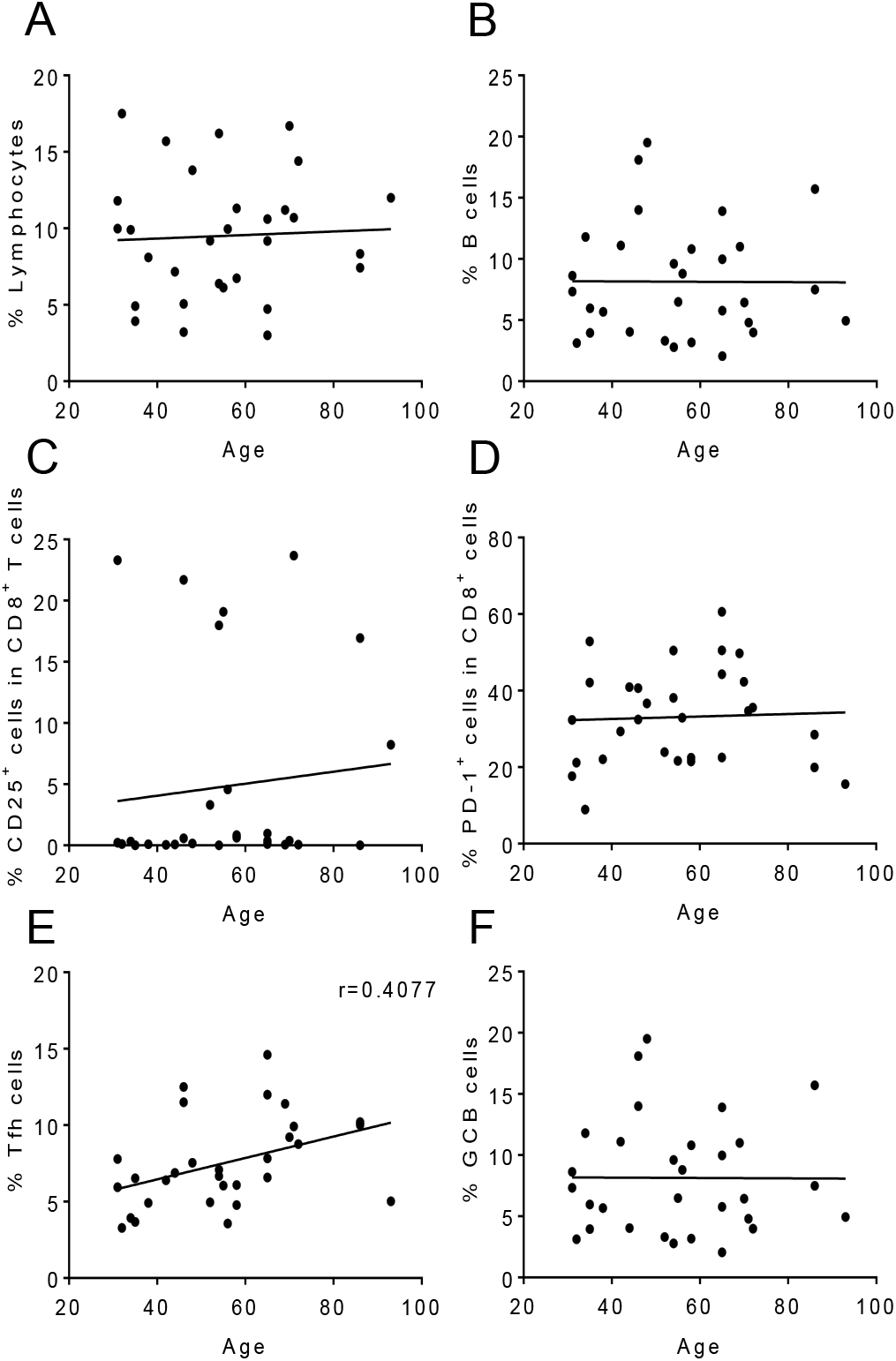
Correlation analysis between functional signatures and patient ages. The correlation analysis between patient age and immune parameters was performed using Pearson’s correlation coefficient. The percentage of total lymphocytes (A), B cells (B), CD8^+^CD25^+^ T cells (C), CD8^+^PD-1^+^ T cells (D), Tfh-like cells (E) and GCB-like cells (F) were correlated to age in COVID-19 patient group. Each dot represents a single patient of COVID-19 or healthy donor. Patients under the age of 15 years were excluded.

## Discussion

SARS-CoV-2 infection is quickly spreading around the world. The patients present typical symptoms of pneumonia, such as dry cough, dyspnea, fever, and bilateral lung infiltrates on imaging [20]. Although the postmortem study revealed intensive inflammation in the patient’s lung, little is known about the immunological features in response to this new virus. In the present study, we have focused on characterizing adaptive immune cell populations and phenotypes in COVID-19 patients.

Lymphopenia was shown in COVID patients from previous studies [20], Epidemiological investigation of coronavirus infection showed that lymphopenia is present in more than 80% of patients, and serious decline is correlated to worse prognosis [21]. However, we did not observe a significant decrease of lymphocyte populations in COVID-19 patients. This finding could be attributed to the fact that most of the patients in this study showed mild symptom besides fever. Interestingly, following the division of patients into those with symptomatic and asymptomatic, we observed a decrease in lymphocyte populations in the symptomatic patients (Data not shown). PD-1 is a marker of exhausted T cells during chronic and acute infections [22, 23]. A number of studies showed that PD-1^+^CD8^+^T cells increased in the peripheral blood of patients with a variety of acute viral infections such as HBV, HIV, and Ebola virus [24, 25]. In our study, the expression of PD-1 was upregulated in both CD4^+^ and CD8^+^ T cells of COVID-19 patients, which may explain the observed reduction in the lymphocyte population.

In response to viral infection, normally both CD4^+^ and CD8^+^ T cells become activated. In COVID-19 patients, we observed a very mild activation in CD4^+^ T cells but stronger activation in CD8^+^ T cells based on CD25 expression. This reflects that CD8^+^ cells are major responders of COVID-19 infection and are consistently activated. It is possible that CD4^+^ T cells may be strongly activated earlier during infection then revert to the quiescent state after providing helper functions, which could explain that lack of detection of a strong activated phenotype. This could be explained by the comparable expression of CD69, an early activation marker in HD and patients. CD4^+^ T cells may indeed be weakly activated in response to the virus, which warrants further studies.

During viral infections, the antigen-specific immune response is executed by Tfh and GCB cells. Tfh cells help B cell differentiate into antigen-specific effector cells to produce high-affinity antibodies and facilitate germinal center formation [19], which are essential for inducing efficient virus clearance. In COVID-19 patients, there was an increase in both Tfh- and GBC-like cells in the blood, reflecting that an antigen-specific response can be activated upon SARS-CoV-2 infection.

Elderly individuals typically exhibit a reduction of the lymphocyte population and weaker ability to defend against viral infection [26]. In our correlation analysis, we did not observe a significant correlation between lymphocyte proportions, effector features, and age. Our data suggest that the specific populations of T and B cells for SARS-CoV-2 are reserved in aged individuals, which need to be proven by further repertoire sequencing analysis of T cell- and B cell-receptors.

In summary, our study shows that SARS-CoV-2 could induce relatively normal adaptive immune response. Most people across different age groups are capable of mobilizing the adaptive immune cells, activating cellular and humoral immunity to defend against the virus with sufficient medical care and anti-viral treatment.

## Materials and Methods

### Ethics statement

This study was approved by the Research Ethics Commission of the Eighth Hospital of Xi’an (20190730-1346) with a waiver of informed consent due to a public health outbreak investigation. All cases were taken from the Eighth Hospital of Xi’an (Xi’an, Shaanxi Province), a designated hospital for the COVID-19 by local authority.

### Patients

From February 18 to March 4, 2020, 18 healthy controls and 38 confirmed COVID-2019 patients were included in the study. Patients were diagnosed and admitted in accordance with the guideline of the national health commission of China. All of 38 patients were confirmed as SARS-CoV-2 infection using the RT-PCR test on throat swab specimens. The median age of the patients was 39.06±4.26 years (Table 1). 23 patients (60.53%) were men, and 15 patients (39.47%) were women (Table 1).

### Flow cytometry analysis

The Abs used in the flow cytometry analysis were as follows: FITC anti-human CD3 (UCHT1), FITC anti-human TCR γ/δ (B1), APC/Cyanine7 anti-human CD4 (OKT4), PerCP/Cyanine5.5 anti-human CD8 (SK1), APC anti-human CD19 (HIB19), APC anti-human CD25 (BC96), PE anti-human CD69 (FN50), PE anti-human CD185 (CXCR5) (J252D4), PE anti-human CD183 (CXCR3) (G025H7), APC anti-human CD279 (PD-1) (EH12.2H7), PE anti-human CD95 (Fas) (DX2), PE anti-human CD127 (A019D5), APC/Cyanine7 anti-human CD45RA (HI100), PE/Cy5 anti-human CD45RO (UCHL1), PE anti-human CD95 (Fas) (DX2), FITC anti-mouse/human GL7 Antigen(GL7), were purchased from Biolegend. Blood cells were stained with Abs in the dark at room temperature for 15 min, and analyzed on a FACSCanto II flow cytometer (BD Biosciences). FlowJo 8 was used for data analysis.

### Statistical analysis

The continuous variable of normal distribution is represented by mean ± standard deviation, the non-normal distribution is represented by median [IQR], and the classified variable is represented by count (percentage). The student’s t test was performed for two group analysis using SPSS 22.0 software. * and ** strands for *P*<0.05 and *P*<0.01, respectively.

## Data Availability

All the data are available in the manuscript.

## ACKNOWLEDGEMENT

This work was supported by grants from Natural Science Foundation of China (No. 81820108017), Natural Science Foundation of China (No. 81771673), and COVID-19 special project of Xi’an Jiaotong University Foundation (xzy032020002). We thank all the doctors, nurses, public health workers, and patients for their contribution against SARS-CoV-2 infection.

## Author contributions

Xiaofeng Yang, Xiaobo Zhou, Lei Lei, Xingzhe Zhang, Dan Zhang analyzed the data and wrote the manuscript. Tongxin Dai, Hongbo Qian, Rui Guo and Yaling Guo collected samples and information. Lin Shi and Yanbin Cheng discussed data analysis. Jinsong Hu performed the FCAS analysis. Baojun Zhang generated the idea, designed the experiment and wrote the manuscript. All authors agree to be responsible for their own part of the work.

**Table 1. Characteristic analysis of COVID-19 patients and healthy donors**

